# Wearable Robotic Ankle Resistance Training Improves Neuromuscular Control and Walking Efficiency in Cerebral Palsy

**DOI:** 10.1101/2020.10.23.20218008

**Authors:** Benjamin C. Conner, Michael H. Schwartz, Zachary F. Lerner

## Abstract

Cerebral palsy (CP) is characterized by deficits in motor function due to reduced neuromuscular control. We leveraged the guiding principles of motor learning theory to design a wearable robotic intervention intended to improve neuromuscular control of the ankle. The goal of this pilot clinical trial was to determine the response to four weeks of exoskeleton ankle resistance therapy (exo-therapy) in children with CP. Five children with CP (12 – 17 years, GMFCS I – II, four males and one female) were recruited for ten, 20-minute sessions of exo-therapy. Surface electromyography, three-dimensional kinematics, and metabolic data were collected at baseline and after training was complete. Changes in neural complexity (via muscle synergy analysis) and metabolic cost were compared to retrospective age- and GMFCS-matched controls who had undergone either single event multi-level orthopedic surgery (SEMLS) or selective dorsal rhizotomies (SDR). Participants displayed decreased co-contraction at the ankle (−29 ± 11%, p = 0.02) and a more typical plantar flexor activation profile (33 ± 13%, p = 0.01), and improvements in neuromuscular control led to a more mechanically-efficient gait pattern (58 ± 34%, p < 0.05) with a reduced metabolic cost of transport (−29 ± 15%, p = 0.02). There were significant increases in neural complexity (5 ± 3%, p = 0.03), where were significantly greater than those seen with SEMLS and SDR (p < 0.01 for both). Ankle exoskeleton resistance therapy shows promise for rapidly improving neuromuscular control for children with CP, and may serve as a meaningful rehabilitative complement to common surgical procedures.

## I. Introduction

The ankle plantar flexor muscles play a key role in mechanical energy recovery while walking, extending the knee joint, preventing excessive ankle dorsiflexion during midstance, modulating center of mass vertical displacement, and providing the single-largest contribution to forward propulsion across all lower-extremity muscle groups [1], [2]. This allows the vertical and horizontal movement of the body’s center of mass to follow an arced, inverted pendular pattern that provides an effective exchange in potential and kinetic energy for maximal efficiency while walking [3]. Activation of the ankle plantar flexor muscles is reduced, less modulated, and often accompanied by co-activation of the antagonist dorsiflexor muscles in a majority of individuals with spastic cerebral palsy (CP) [4], a movement disorder arising from injury to the brain during infancy [5]. These muscle activation characteristics likely contribute directly or indirectly to reduced energy exchange [6], elevated metabolic cost of transport [7], and lower levels of physical activity [8] in this patient population.

Impaired neuromuscular control in children with CP may, in part, be explained by deficits in cortical organization [9]. It has been shown that the variance in muscle activity accounted for by one muscle synergy is associated with the level of motor control complexity [10]. Children with CP have greater variance accounted for by one muscle synergy while walking compared to typically developing peers, leading to the conclusion that these children use a simplified control strategy [11]. This measure of motor control in children with CP plays a significant role in explaining treatment outcomes for this population [12].

The current standard of care for children with CP has not been successful at improving neuromuscular control in this population. Orthopedic surgery and selective dorsal rhizotomies, while a critical component in the care of musculoskeletal health and management of spasticity for individuals with CP [13], may have limited influence on the neuromuscular control of movement [14]. Lower-limb orthoses, which serve to mitigate misalignment and joint contractures [5], are not specifically designed to train motor control performance. Traditional physical therapy has shown limited evidence of improving motor function and walking ability [15], [16]. Gait training with partial body weight support can be help individuals with more severe impairment achieve task-specific, repetitive practice, but prolonged unloading over time may lead to muscle atrophy [17] and could impact afferent signaling from load receptors that are important for muscle activation timing [18]. Functional electrical stimulation is a bottom-up approach that has failed to produce lasting changes in neuromuscular control or gain traction as an effective tool for treating CP [19]. There is an apparent gap in our ability to effectively address deficits in the neuromuscular control of walking for individuals with CP.

The primary goal of this pilot clinical trial was to develop and evaluate a task-specific gait re-training intervention to improve neuromuscular control and walking efficiency in adolescents with CP. This work builds on our previous validation of an adaptive ankle resistance control scheme implemented on a light-weight wearable exoskeleton to foster increased volitional engagement of the ankle plantar flexor muscles [20]. We hypothesized that repeated neuromuscular gait re-training with adaptive ankle resistance that responds directly to user input would improve the ankle plantar flexor muscle activation profile and decrease co-contraction across the ankle joint. We further hypothesized that improvement in plantar flexor activation and reduced co-contraction would lead to a more mechanically and metabolically efficient gait pattern. To test these hypotheses, we recruited five children with spastic CP for a four-week training intervention with wearable adaptive resistance. Our secondary goal was to determine how ankle exoskeleton resistance training modulated motor control complexity, and to compare this modulation with changes resulting from the most common invasive interventions used in the treatment of CP – single event multi-level surgery (SEMLS) and selective dorsal rhizotomy (SDR). We hypothesized that anticipated improvement in motor control and energetic efficiency following adaptive ankle resistance would be reflected by cortical reorganization for a more complex control strategy, as indicated by muscle synergy analysis. In addition, we completed a retrospective analysis of matched controls to test the hypothesis that adaptive resistance would result in similar or greater improvements compared to SEMLS or SDR.

### II. METHODS

This study was approved by the Northern Arizona University Institutional Review Board (#986744). The protocol was completed at the Northern Arizona University – Phoenix Biomedical Campus (Phoenix, AZ) and registered at ClinicalTrials.gov (NCT04119063). Informed written consent was provided by a parent or legal guardian for each participant after the nature and possible consequences of the study was explained; participants provided verbal assent.

### A. Participants

We recruited 6 individuals diagnosed with hemiplegic or diplegic spastic CP based on the following inclusion criteria: Gross Motor Function Classification System (GMFCS) levels I – II, the ability to walk with or without a walker for at least six minutes, age between 10 – 21 years, and the ability to follow simple directions. Individuals were excluded from the study if they had orthopedic surgery within the past six months, or any conditions that would prevent safe participation. Data from five participants are reported in this study due to equipment failure and the resulting absence of experimental biomechanics data needed for this analysis (Table 1).

**TABLE I.** PARTICIPANT CHARACTERISTICS.

| Participant | Gender | Age range (y) | GMFCS<sup>a</sup> | Gait type<sup>b</sup> |
| --- | --- | --- | --- | --- |
| P1 | M | 13-16 | II | Crouch |
| P2 | M | 13-16 | II | Crouch |
| P3 | M | 13-16 | I | Crouch |
| P4 | F | 13-16 | I | Stiff knee |
| P5 | M | 10-13 | II | Crouch |

<sup>a</sup>GMFCS: Gross Motor Function Classification System
<sup>b</sup>Gait type: ‘Crouch’ gait defined by those parameters set by Gage et al [2]

### B. Wearable Resistance Platform

Our ankle exoskeleton resistance therapy (exo-therapy) utilized a custom battery-powered lower limb exoskeleton that provided resistance to ankle plantar flexion (Fig. 1a). The device had an actuation and control assembly that was worn at the waist, and ankle assemblies worn bilaterally. Both hemiplegic and diplegic participants wore a bilateral exoskeleton for consistency. The actuation and control assembly consisted of two high-powered DC motors that actuated a steel cable attached to a pulley aligned with each ankle joint in the sagittal plane. Each ankle assembly was constructed from carbon fiber and consisted of a foot plate with embedded force sensors, a calf cuff, pulley, and torque sensor aligned with the ankle joint. The torque sensors were used for low-level proportional-derivative motor control. Detection of gait events and real-time estimates of ankle moment from the embedded force sensors allowed the device to provide proportional resistance to ankle plantar flexion [20]. An onboard microcontroller with Bluetooth input allowed for remote control via a custom MATLAB graphical user interface and visualization of the estimated ankle movement and exoskeleton torque. This allowed the research team to assess, in real-time, user performance with the device so that active engagement could be maximized. Additional details on the device can be found in earlier work [20].

**Fig. 1.**
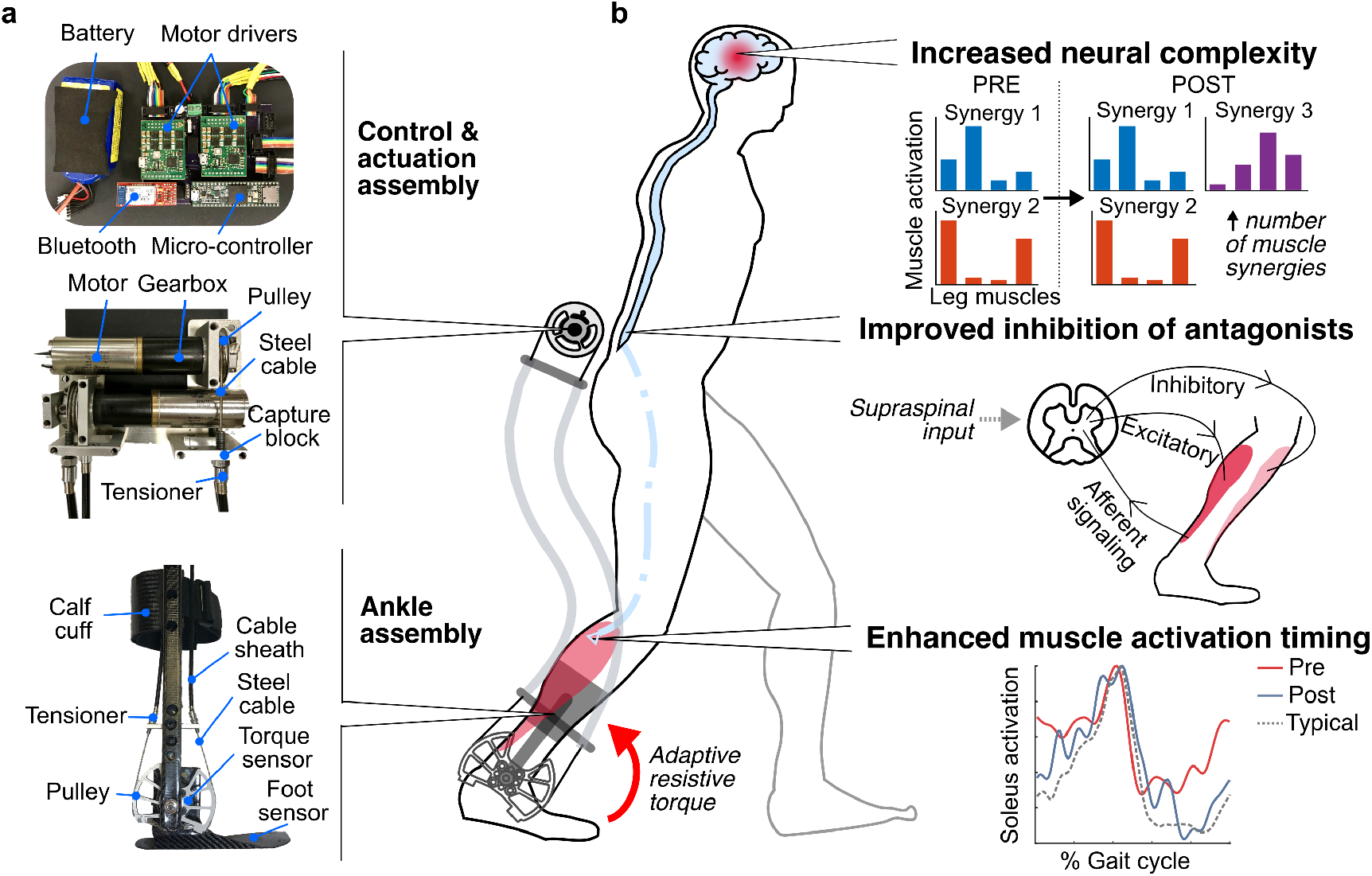
Wearable adaptive resistance training. a) Pictures of the exoskeleton used to deliver adaptive ankle resistance b) Schematic depiction of the proposed mechanisms underlying the neuromuscular response to wearable adaptive resistance training, including increased neural complexity (as indicated by an increase in the number of muscle synergies recruited for walking), improved inhibition of antagonist muscles about the ankle, and enhanced muscle activation timing of the plantar flexor muscles for more typical activation timing (experimental data from P1).

### C. Pre- and Post-Assessment

On the first visit, participants were evaluated by a licensed physical therapist to determine physical characteristics and GMFCS level. Using the physical therapist’s assessment and input from parents/guardian, each participant’s more- and less-affected lower-limbs were determined. For the pre- and post-assessments, wireless surface electromyography (EMG) sensors (Noraxon, Scottsdale, AZ; 1000 Hz) were placed bilaterally on the soleus (SOL), tibialis anterior (TA), vastus lateralis (VL), and semitendinosus (ST) according to SENIAM recommendations that uses bony landmarks for consistency between visits [21]. Participants were then outfitted with 38 reflective markers for measuring 3D kinematics of the lower extremities, pelvis, and trunk using eight motion capture cameras (Vicon, Denver, CO; 100 Hz). Participants walked on a treadmill at a self-selected speed while being monitored for safety by a laboratory technician. Preferred walking speed was identified by asking participants to choose a speed that they would normally walk at while at school or home. The treadmill speed was then increased and decreased to confirm preferred walking speed. Three-dimensional kinematics and EMG activation levels were recorded for 30 seconds of walking.

Oxygen and carbon dioxide levels were assessed using a metabolic measurement system (TrueOne 2400, Parvo Medics, Salt Lake City, UT, USA). Metabolic measurements were taken while participants stood for two minutes, followed by six minutes of walking at their preferred treadmill speed, or until oxygen consumption levels stabilized. During the post-assessment, if the participant’s preferred speed differed from his or her preferred speed during the pre-assessment, metabolic measurements were taken at the pre-assessment preferred speed so that pre- and post-assessment speeds were matched.

### D. Exoskeleton Ankle Resistance Therapy

Training consisted of ten, 20-minute treadmill walking sessions with ankle resistance at each participant’s preferred speed. One to two rest periods were provided during each session, depending on participant preference. The nominal resistance setpoint was initialized between 0.025 – 0.075 Nm/kg, which represents the maximal amount of resistance that would be applied during a gait cycle (i.e., the amount of resistance applied when the peak biological ankle moment was reached, which typically occurred at push-off). Real-time visualization of the estimated biological ankle moment allowed the research team to assess how well participants were reaching the prescribed level of resistance. Participants were instructed to focus on engaging their ankle plantar-flexor muscles, with an emphasis on their more-affected limb. Verbal coaching was provided as needed. Following the completion of each session, participants were asked to rank their level of soreness on the following scale: none, mild, moderate, severe, and extremely severe. If participants had both 1) a perceived level of soreness between none and moderate, and 2) reached their prescribed level of resistance for more than 50% of the training session, then the prescribed resistance level was increased by 0.5 – 1 Nm for the next training session.

### E. Data Processing & Statistical Analysis

A generic OpenSim musculoskeletal model was scaled to the anthropometrics of each participant [22]. Joint angles across 10 continuous gait cycles were calculated in OpenSim via inverse kinematics (pre- and post-musculoskeletal gait model videos can be found in Supplemental Video S1). Participant EMG signals were bandpass filtered (4th order Butterworth, 20 - 400 Hz band-pass cutoff), rectified, and low-pass filtered (4th order Butterworth, 10 Hz low-pass cutoff), and then time normalized from 0 – 100% of the gait cycle [23]. The EMG curves were then averaged over the same 10 gait cycles and normalized to the maximal activity recorded during walking within a visit for each respective muscle [24]. For consistency across participants with diplegia and hemiplegia, all EMG measures were assessed for the more-affected limb only.

Co-contraction between the soleus and tibialis anterior was calculated using a co-contraction index (CCI) [25], which captures the temporal and magnitude components of an EMG signal [26], as in (1):

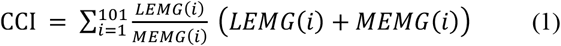

where *i* represents the individual time points of the time-normalized gait cycle (0 – 100%, or 101 total data points), *LEMG* represents the normalized magnitude of the less active muscle at time point *i*, and *MEMG* represents the normalized magnitude of the more active muscle at time point *i*.

We assessed the relationship between the soleus EMG profile for our participants and the typical (control) soleus EMG profile from unimpaired individuals by calculating the Pearson product moment correlation coefficient (*R*) [27]. We used a publicly available data set for the typical soleus EMG curve from unimpaired individuals walking at a non-dimensional speed of 0.25 [28], which was very close to the average non-dimensional speed of our participants (0.21); non-dimensional walking speed accounts for leg length and was calculated by dividing walking speed by the square root of gravity multiplied by leg length [29].

The variance in muscle activity accounted for by one muscle synergy (VAF_1_) was calculated using non-negative matrix factorization (NNMF) [30] [11], as in (2):

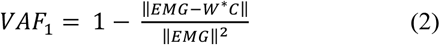

where *EMG* represents a matrix containing the normalized and averaged EMG data recorded for the SOL, TA, VL, and ST; *W* represents the relative activation level of each muscle in a synergy and is a 4 x 1 matrix, with a separate row for each muscle; *C* represents the activation level of a synergy over the gait cycle and is a 1 x 101 matrix, with a separate column for each time point. Using NNMF with random initial values of *W* and *C*, an iterative optimization was run to minimize the error in EMG data calculated by *W*C*. The resulting values for *W* and *C* were then used to calculate VAF_1_. VAF_1_ was calculated for both the more-affected and less-affected limb, and then averaged between limbs for a single VAF_1_ value.

The mechanical efficiency of each individual’s pre- and post-training gait patterns was calculated using energy recovery analysis (Fig. 4) [6]. Center of mass mechanical energy recovery is significantly lower in children with CP [6], which may contribute to the higher metabolic cost of walking in this population [31]. This measure was quantified as the exchange between kinetic and potential energy of the center of mass (COM) movement (5) by considering the external work (W_ext_) (3) on the COM with perfect exchange, and external work on the COM with no exchange (W_ne_) (4) [6]:

**Fig. 2.**
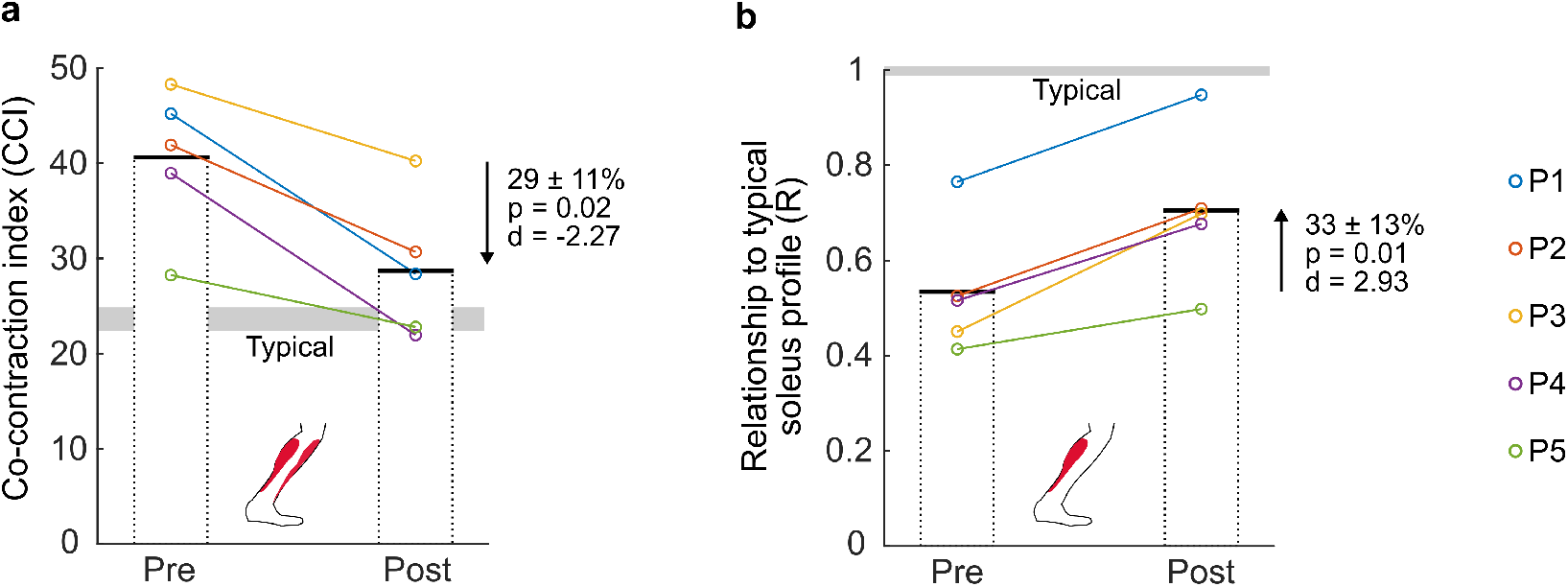
Neuromuscular outcome measures. Individual (color-coded circles) and group mean (bars) pre- and post-exo-therapy neuromuscular variables, including a) co-contraction index (CCI) between the soleus and tibialis anterior; b) the relationship of the experimental soleus activation profile to a typical activation profile at a non-dimensional speed of 0.25; ‘Typical’ neuromuscular values (a,b) were calculated using a publicly available dataset [28] of individuals walking at a non-dimensional speed of 0.25.

**Fig. 3.**
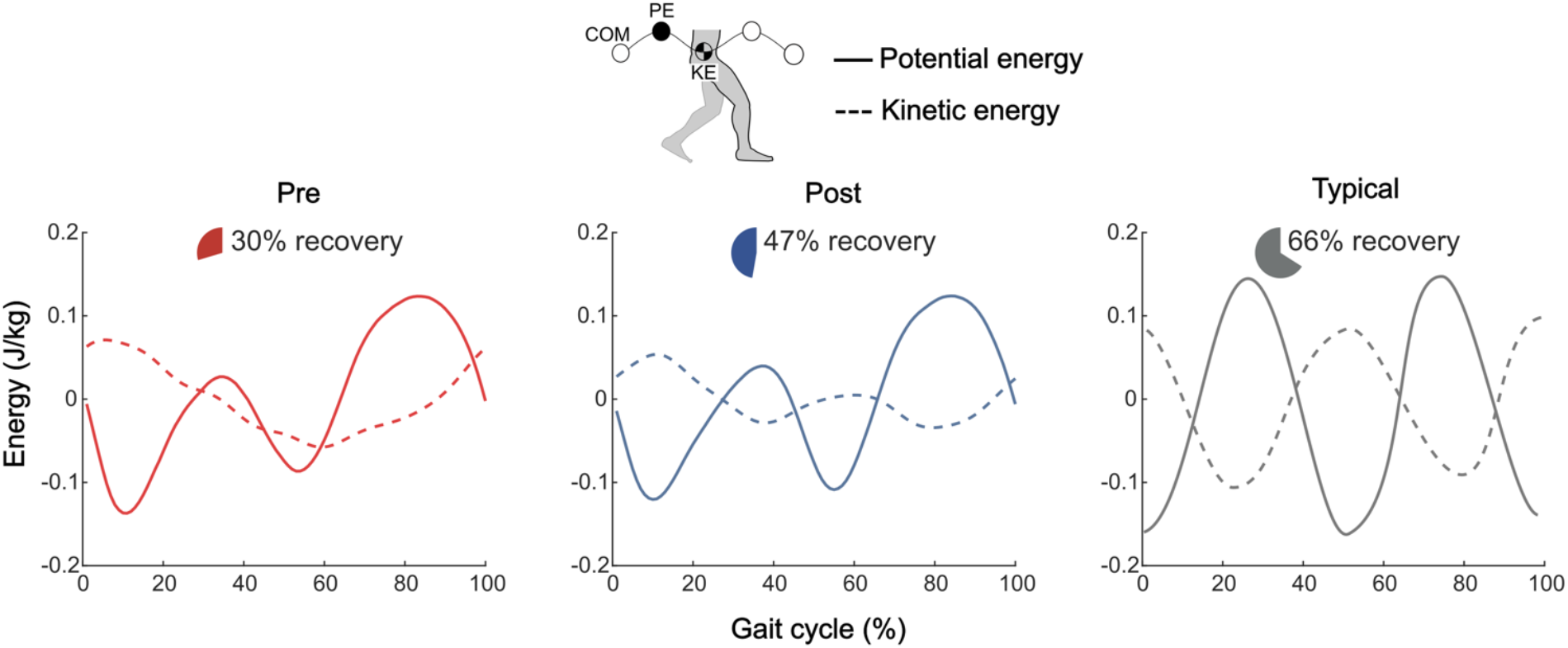
Center of mass mechanical efficiency. a) Group average pre-exo-therapy (red), group average post-exo-therapy (blue), and typical unimpaired (gray) potential energy (PE, solid lines) and kinetic energy (KE, dashed lines) curves. Pie charts indicate the group average center of mass (COM) energy exchange recovery percentage; For efficient energy exchange and increased recovery, potential and kinetic energies should be 180° out of phase with equal and opposite amplitudes. The typical, unimpaired curve was adopted from Bennett et al [6].

**Fig. 4.**
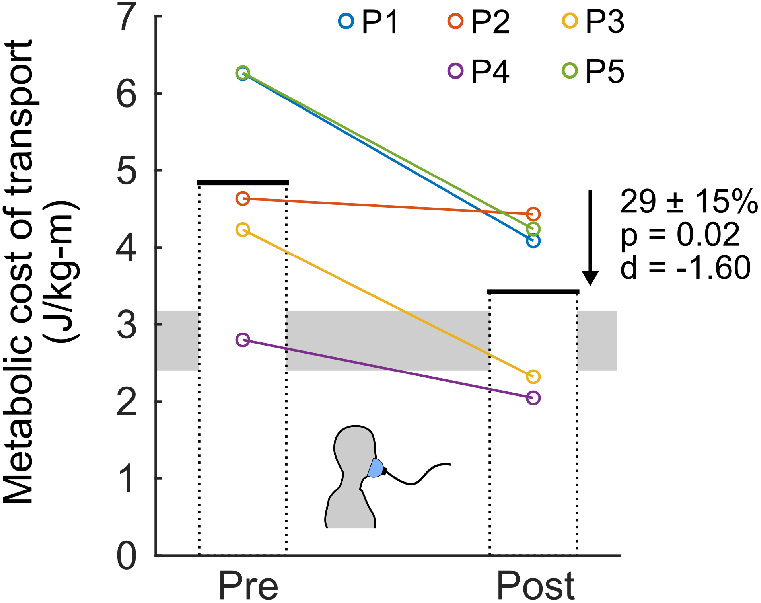
Metabolic cost. Individual (color-coded circles) and group mean (bars) pre- and post-exo-therapy metabolic cost of transport, representing the body-mass-normalized metabolic energy required to walk a unit distance. Both pre- and post-measures were collected at the pre-exo-therapy preferred walking speed on the treadmill.

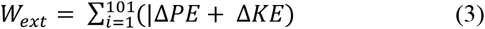

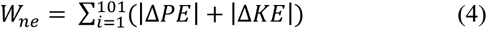

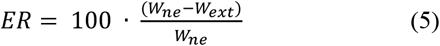

where *i* represents the individual time points of the time-normalized gait cycle (0 – 100%, or 101 total data points), *ΔPE* represents the change in potential energy of the COM between *i* and *i* + 1, and *ΔKE* represents the change in kinetic energy of the COM between *i* and *i* + 1.

Metabolic cost of transport was calculated using participants’ expired gas data (TrueOne 2400, Parvo Medics, Salt Lake City, UT, USA). First, steady state regions for both quiet standing and walking data were determined by Kendall’s tau-b approach [32], which has been shown to contribute to a five-fold reduction in variability of measured oxygen costs while walking for individuals with CP [32]. Metabolic cost of each region was calculated using Brockway’s standard equation [33], and a net metabolic cost (W) was determined by subtracting the metabolic cost of quiet standing from the metabolic cost of walking. Net metabolic cost was then divided by body mass (kg) and walking speed (m/s) for a final measure of body-mass-normalized metabolic energy required to walk a unit distance (i.e., metabolic cost of transport, J/kg-m).

Our primary outcome measures included CCI, relationship of experimental soleus EMG profile to a typical profile, energy recovery, and metabolic cost of transport. All primary outcome variables were assessed at matched speeds (i.e., pre vs. post-training, both at the initial preferred speed). Our secondary outcomes included VAF_1_ and a matched-comparison of changes in VAF_1_ to SEMLS and SDR. Changes in VAF_1_ from pre- to post-exo-therapy were also evaluated at matched speeds.

We assessed all outcome measure data for normality and the presence of outliers. Normality was tested using the Kolmogorov-Smirnov test with small sample Lilliefors correction [34]. Outliers were defined as any data point below the first quartile or above the third quartile of the 1.5 interquartile range [35]. Datum falling within this outlier definition were removed for any statistical analyses. We evaluated our primary outcome measures using two-tailed paired t-tests with Holm-Bonferroni correction to account for multiple comparisons. Significance level was set at α < 0.05. Cohen’s d (d) was used to calculate effect size, where 0.2 was considered a small effect, 0.5 a medium effect, and 0.8 a large effect [36].

We assessed our secondary outcome measure of pre- vs. post-training VAF_1_ using a two-tailed paired t-test. We also assessed the relationship between VAF_1_ and CCI by calculating the Pearson product moment correlation coefficient (R) using combined pre and post data. Additionally, we retrospectively matched our participants by age and GMFCS level to patients who underwent SEMLS or SDR at Gillette Children’s Specialty Healthcare (St. Paul, MN, USA) from 2005 – 2018. The three most-similar age- and GMFCS-matched controls were assigned to each study participant for both SELMS and SDR (i.e., total of six matched controls/participant; Table 2; see Supplemental Table S1 for individual matched-control characteristics). Two-tailed, independent t-tests were run to assess differences in the change in VAF_1_ between exo-therapy vs. SEMLS and SDR. Significance level was set at α < 0.05.

**TABLE II.** MATCHED-CONTROL CHARACTERISTICS.

| Intervention type<sup>a</sup> | Average age (y:m) | GMFCS<sup>b</sup> |
| --- | --- | --- |
| Exo-therapy (n = 5) | 14:9 | I (n = 2), II (n = 3) |
| SEMLS (n = 15) | 13:6 | I (n = 6), II (n = 9) |
| SDR (n = 15) | 9:11 | I (n = 6), II (n = 9) |

<sup>a</sup>Intervention type: Exoskeleton ankle resistance therapy (Exo-therapy), Single event multi-level orthopedic surgery (SEMLS), selective dorsal rhizotomy (SDR). <sup>b</sup>GMFCS: Gross Motor Function Classification System

## III. RESULTS

All participants tolerated progressive increases in resistance. The level of resistance reached by the final training visit ranged from 0.14 – 0.27 Nm/kg. Post-training soreness ranged from none – severe across all sessions.

Following exo-therapy, more-affected limb co-contraction between the soleus and tibialis anterior decreased by 29 ± 11% (p = 0.02, d = -2.27; Fig. 2a) and the relationship (R) between experimental and the typical soleus curve increased by 33 ± 13% (p = 0.01, d = 2.93; Fig. 2b; see Supplemental Figure S1 for individual participant EMG curves) during walking at the same speed.

Whole-body center of mass potential-kinetic energy exchange recovery increased by 58 ± 34% (p = 0.04, d = 1.57; Fig. 3; see Supplemental Figure S2 for individual energy curves) when walking at the same preferred speed, with a pre-assessment average of 30% and a post-assessment average of 47% (typically developing average is 66%).

Metabolic cost of transport decreased by 29 ± 15% (p = 0.02, d = -1.60; Fig. 4, see Supplemental Figure S3 for individual metabolic curves) when walking at the same speed.

The variance in muscle activity of the soleus, tibialis anterior, vastus lateralis, and semitendinosus explained by one muscle synergy decreased by 5 ± 3% (p = 0.03, d = -1.52; Fig. 5a). There was a significant relationship between participants’ VAF_1_ and CCI when combining both pre and post data (R = 0.85, p = 0.002; Fig. 6). When compared to age- and GMFCS-matched control data (Table 2), exo-therapy resulted in greater reduction in VAF_1_ than both SEMLS (p = 0.005) and SDR procedures (p = 0.008) (Fig. 5b).

**Fig. 5.**
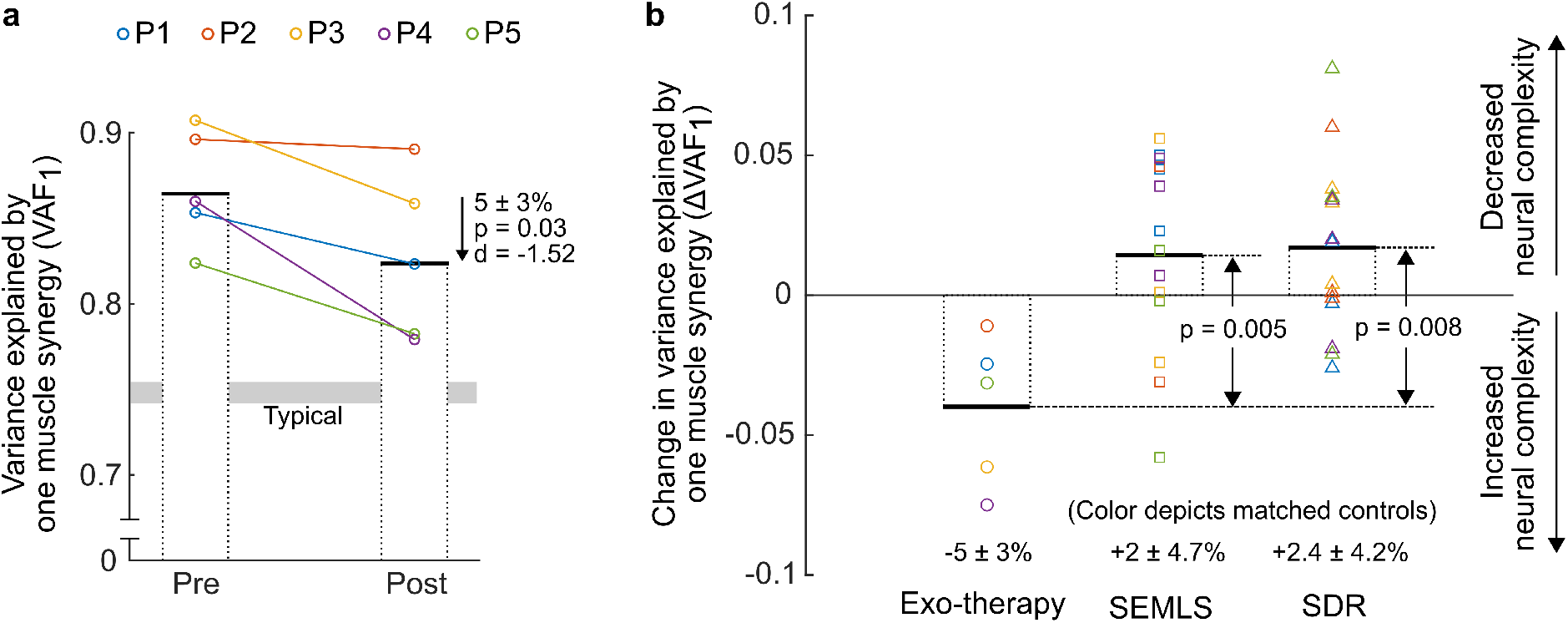
Change in the neural complexity. a) Pre- and post-exo-therapy individual (color-coded circles) and group mean (bars) variance of muscle activity explained by one muscle synergy (VAF_1_); b) Retrospective comparison of change in VAF_1_ (ΔVAF_1_) for three interventions: exoskeleton ankle resistance therapy (Exo-therapy, circles), single event multi-level orthopedic surgery (SEMLS, squares), and select dorsal rhizotomy (SDR, triangles). Both SEMLS and SDR have three, age- and GMFCS-matched controls for each study participant with the same corresponding color. Increases in VAF_1_ (i.e., positive change) represent a decrease in neural complexity, while decreases in VAF_1_ (i.e., negative change) represent an increase in neural complexity.

**Fig. 6.**
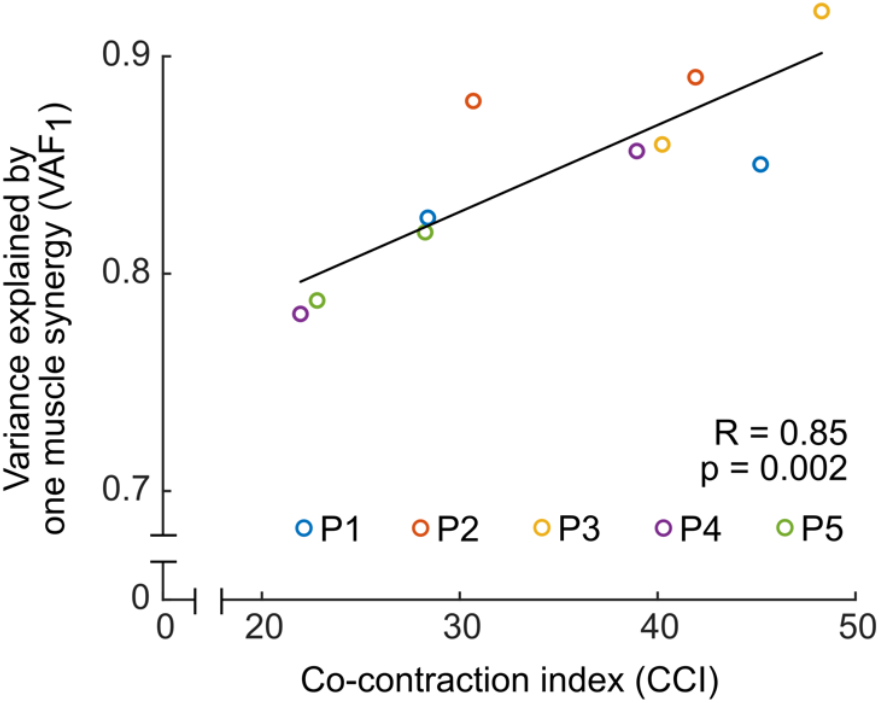
Relationship between muscle synergy and co-contraction. Combined pre- and post-intervention muscle activity variance explained by one synergy (VAF_1_) plotted against combined pre- and post-intervention co-contraction index (CCI) between the soleus and tibialis anterior.

## IV. DISCUSSION

The results of this study support our central hypotheses that exo-therapy would improve plantar flexor activity during walking in children with CP (Fig. 2a,2b), translating to a more mechanically (Fig. 3) and metabolically (Fig. 4) efficient gait pattern. In children with CP, ankle power is significantly reduced [37], potentially due to ineffective muscle activation profiles in which agonist activity is partially negated by antagonist activity. In our prior validation study of the resistance controller, we observed a significant decrease in ankle co-contraction when individuals walked with resistance [20]. The present study found that there was a carry-over of this desirable reduction in co-contraction to walking without the device following exo-therapy. We theorize that reductions in co-contraction may be due to improvement in reciprocal inhibition (Fig. 2a), which is impaired in children with CP [38]. We also observed a significant increase in the similarity of our participants’ more-affected limb soleus muscle activity profiles compared to the activation pattern from unimpaired individuals (Fig. 2b), reflecting improved activation timing. Improvements in neuromuscular control appear likely to have produced more mechanically (Fig. 3) and metabolically (Fig. 4) efficient gait patterns. Metabolic cost of walking is inversely related to physical activity levels for children with CP [39], signifying promising clinical relevance of our intervention and findings.

We found a modest, but significant reduction in the variance in muscle activity that can be explained by one muscle synergy (VAF_1_) following exo-therapy (Fig. 5a), which suggests the potential for cortical reorganization. We also observed a significant positive relationship between this measure of selective motor control and co-contraction of the agonist and antagonist muscles about the ankle (Fig. 6). This supports the theory that decreased complexity of motor control (i.e., larger VAF_1_) results in greater muscle co-activation in children with CP. Smaller VAF_1_ in children with CP has been shown to be a strong predictor of positive treatment outcomes, independent of the treatment [12]. A rehabilitative tool that is able to lower VAF_1_, therefore, could improve treatment outcomes from SEMLS or SDR for children with CP [40]. Importantly, we found that the increased neural complexity with exo-therapy was significantly greater than the changes seen with SEMLS or SDR (Fig. 5b). This finding supports the use of exo-therapy to complement rehabilitation before or after these common procedures, whereby doing so could induce neuromuscular changes not previously possible with surgery alone.

Exo-therapy was designed to capitalize on the core principles of motor learning, including task-specificity, repetition, and active engagement [41]. Our wearable system allowed participants to train within the task-specific, functional context of walking, and achieve repetitive, high-volume practice. Applying resistance to plantar flexion necessitated active user engagement to maintain speed on the treadmill. Additionally, the adaptive nature of the proportional joint-moment control scheme made it so that resistance was immediately responsive to user input, creating an experience that fostered active engagement.

Afferent signals from load receptors play a critical role in muscle activation timing while walking [42], and the phase-specific resistance of this intervention may have served as a supraphysiologic signal for the plantar flexors to fire at the appropriate time. Motor control theory also dictates that the modular recruitment of muscles (i.e., muscle synergies) become more specific with greater biomechanical constraints [43]. With resistance as a new biomechanical constraint while walking, there may have been a demand for greater precision in motor control by the participants, necessitating increased neural complexity as indicated by the significant changes in VAF_1_. These features of exo-therapy may have worked together to facilitate rapid motor learning for improved neuromuscular control.

The findings from this study are complemented by the potential accessibility of exo-therapy. The platform for exo-therapy is relatively inexpensive and could be readily translated to the clinic or home. In such environments, this technology holds potential to increase the dose of precision therapy by a considerable degree over standard of care. By exploiting core principles of motor learning, exo-therapy is a promising intervention to facilitate the treatment of walking disability across the broad spectrum of neurological disorders beyond individuals with CP.

This pilot study had several limitations. First, we did not include a direct control group to isolate the effects of exo-therapy independent of structured treadmill walking. However, the 20 minutes of training completed by our participants during each visit was likely not a significant departure from their typical daily walking volume. In addition, previous studies on standard treadmill training have not demonstrated robust evidence of effectiveness, resulting in a weak clinical recommendation [16]. To the best of our knowledge, no prior studies of treadmill-only training in CP have reported similar findings to those reported here. Furthermore, while a direct control group was beyond the scope of, and resources available to, this study, we compared the changes in muscle synergy from exo-therapy to age- and GMFCS-matched control data for individuals with CP who underwent SEMLS or SDR. A second limitation of this pilot work was the small sample size (n = 5). This sample size, however, closely matches those of similar pilot studies [24], [44]–[46], and will form the basis for larger, randomized controlled trials. In addition, we used a rigorous statistical analysis, including corrections for multiple comparisons, and reported large to very large effect sizes that demonstrate the consistency of our findings. Finally, all training occurred on a treadmill, which may lack the same ecological validity as overground training for transferring improvements to real world performance. Future studies will explore the feasibility and outcomes of overground training with exo-therapy.

The findings from this pilot clinical trial indicate the unique ability of exo-therapy to improve neuromuscular function of the ankle musculature and enable a more efficient gait pattern in children with CP. We observed improved soleus activation timing and coordination, improved energy transfer and reduced metabolic cost, and increased complexity of neuromuscular control. This novel training modality could supplement the current standard of care for individuals with CP, offering increased access to targeted neuromuscular rehabilitation.

## Data Availability

All data referred to in this manuscript is available by request from the corresponding author (ZFL).

https://biomech.nau.edu/index.php/resources/

## Acknowledgment

The authors would like to thank Nushka Remec, Emily Frank, Elizabeth Orum, and Jason Luque for their assistance with data collections, and James Babers and Leah Liebelt for their assistance with device manufacturing. The authors would also like to thank the participants and their families for their involvement in the study.

## Supplementary Material

Video S1. Pre- and post-intervention scaled musculoskeletal models in OpenSim at preferred walking speed.

Table S1. Individual matched-control participant characteristics.

Figure S1. Individual soleus activation curves.

Figure S2. Individual energy recovery curves.

Figure S3. Individual metabolic cost curves.

