## Supplementary material for "Wearable Robotic Ankle Resistance Training Improves Neuromuscular Control and Walking Efficiency in Cerebral Palsy": Figure S1

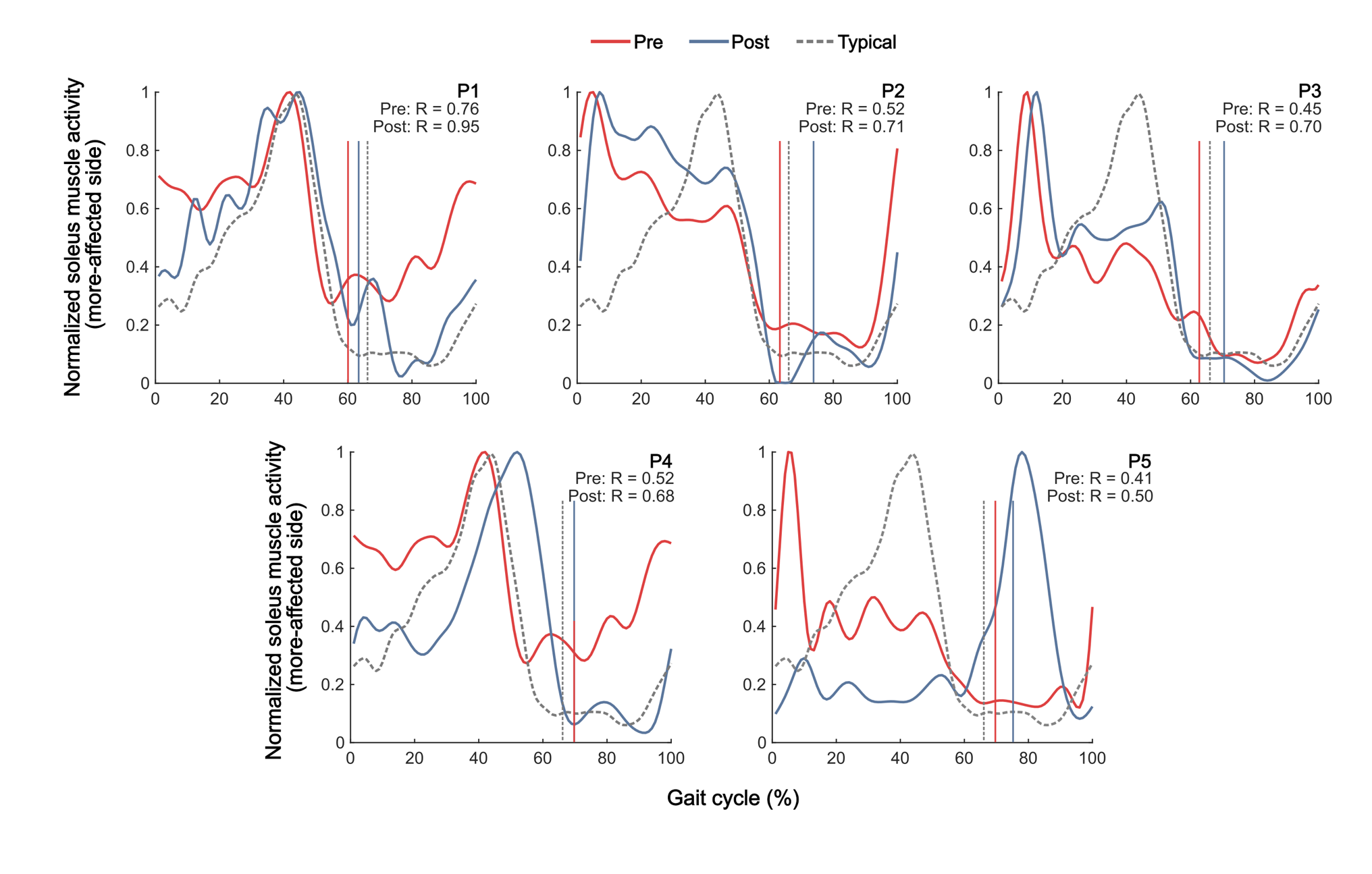

**Figure S1**. **Individual soleus activation curves**. Average, normalized soleus activation curves pre (red) and post (blue) training, both at the same preferred walking speed, compared to a typical soleus activation profile (gray, dashed). Vertical lines indicate pre (solid red), post (solid blue), and typical (dashed gray) toe offs, and “R” indicates the Pearson product moment correlation coefficient between the experimental and typical activation profiles. Experimental curves were normalized to peak activation levels and averaged over ten gait cycles.
