## Supplementary material for "Wearable Robotic Ankle Resistance Training Improves Neuromuscular Control and Walking Efficiency in Cerebral Palsy": Figure S2

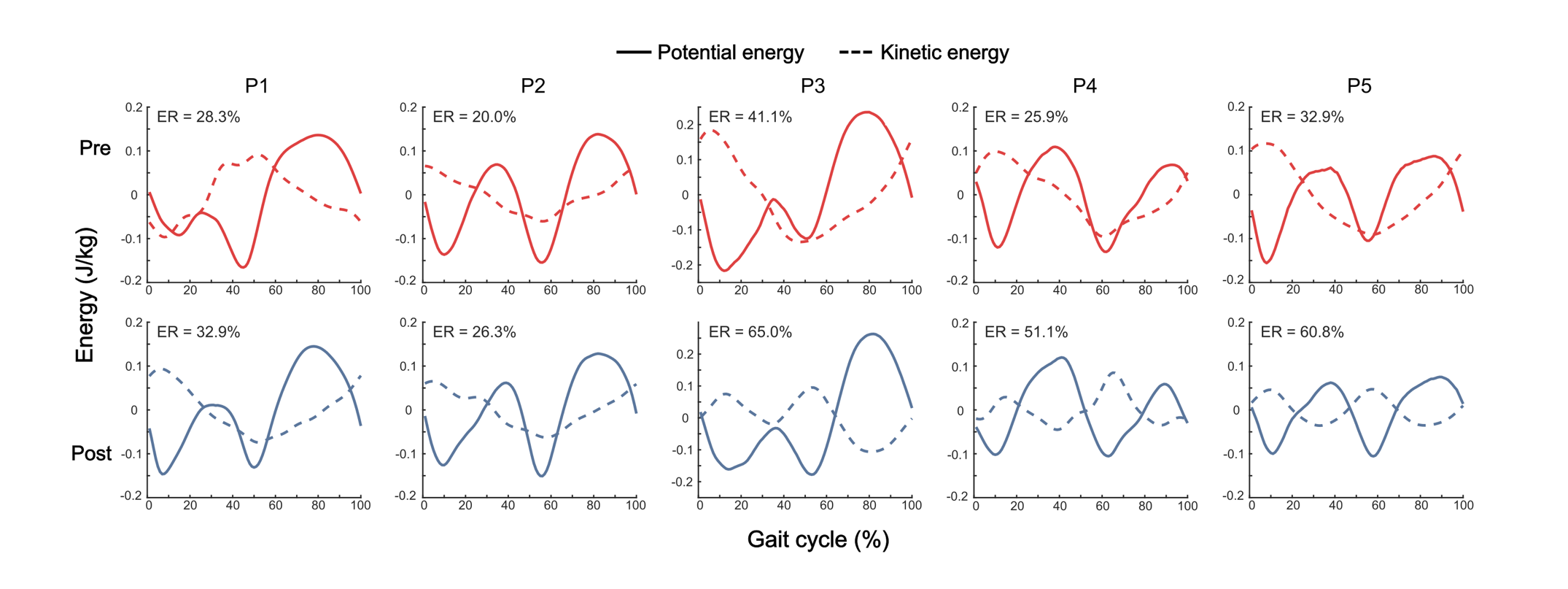

**Figure S2. Individual energy recovery curves.** Pre and post potential energy (solid lines) and kinetic energy (dashed lines) curves at the same preferred walking speed, plotted as variations about their respective means, and individual pre and post energy recovery percentages.
