## Supplementary material for "Wearable Robotic Ankle Resistance Training Improves Neuromuscular Control and Walking Efficiency in Cerebral Palsy": Figure S3

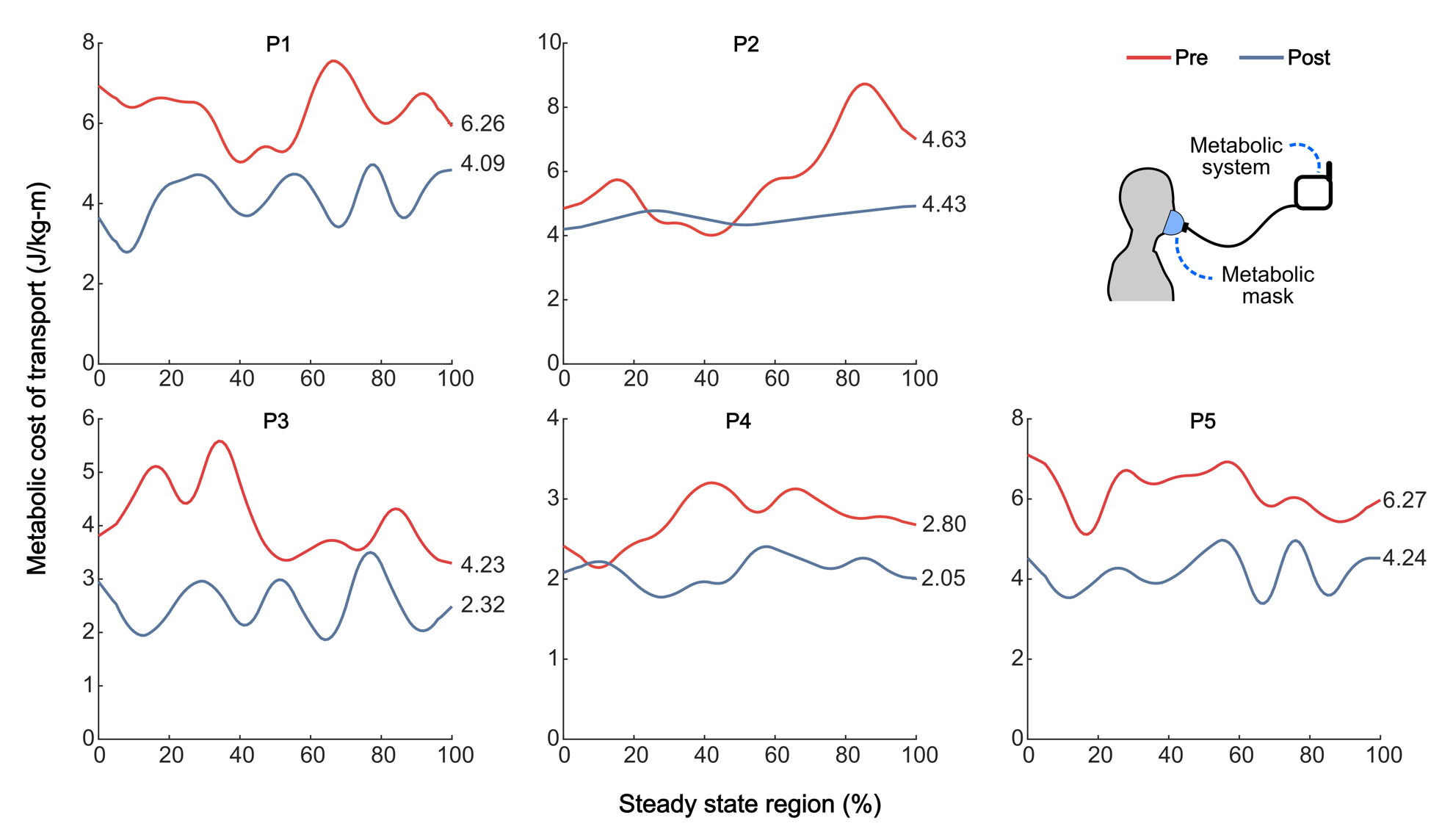

**Figure S3**. **Individual metabolic cost curves**. Pre- (red) and post-exo-therapy (blue) metabolic cost of transport steady state regions for each participant, representing the body-mass-normalized metabolic energy required to walk a unit distance. Values to the right of each curve indicate the average of each steady state region.
