## Supplementary material for "Wearable Robotic Ankle Resistance Training Improves Neuromuscular Control and Walking Efficiency in Cerebral Palsy": Table S1

**Table S1. Individual matched-control participant characteristics**

| **Participant** | **Matched**  **control** | **Control**  **gender** | **Control age range (y)** | **Control**  **GMFCS level^a^** | **Intervention type^b^** |
| --- | --- | --- | --- | --- | --- |
| **P1** | C1 | M | 13-16 | II | SEMLS |
|  | C2 | M | 16-19 | II | SEMLS |
|  | C3 | F | 16-19 | II | SEMLS |
|  | C4 | M | 13-16 | II | SDR |
|  | C5 | M | 10-13 | II | SDR |
|  | C6 | F | 10-13 | II | SDR |
| **P2** | C7 | F | 10-13 | II | SEMLS |
|  | C8 | F | 10-13 | II | SEMLS |
|  | C9 | M | 13-16 | II | SEMLS |
|  | C10 | F | 10-13 | II | SDR |
|  | C11 | M | 10-13 | II | SDR |
|  | C12 | M | 10-13 | II | SDR |
| **P3** | C13 | M | 13-16 | I | SEMLS |
|  | C14 | M | 13-16 | I | SEMLS |
|  | C15 | F | 13-16 | I | SEMLS |
|  | C16 | F | 10-13 | I | SDR |
|  | C17 | M | 10-13 | I | SDR |
|  | C18 | F | 10-13 | I | SDR |
| **P4** | C19 | M | 10-13 | I | SEMLS |
|  | C20 | M | 10-13 | I | SEMLS |
|  | C21 | F | 10-13 | I | SEMLS |
|  | C22 | M | 7-10 | I | SDR |
|  | C23 | F | 7-10 | I | SDR |
|  | C24 | M | 7-10 | I | SDR |
| **P5** | C25 | F | 10-13 | II | SEMLS |
|  | C26 | F | 10-13 | II | SEMLS |
|  | C27 | F | 10-13 | II | SEMLS |
|  | C28 | F | 7-10 | II | SDR |
|  | C29 | M | 7-10 | II | SDR |
|  | C30 | M | 7-10 | II | SDR |

^a^GMFCS: Gross Motor Function Classification System.

^b^Intervention type: Single event multi-level orthopedic surgery (SEMLS), selective dorsal rhizotomy (SDR).
